# Do children with disabilities have the same opportunities to play as children without disabilities? Evidence from the Multiple Indicator Cluster Surveys in 38 low and middle-income countries

**DOI:** 10.1101/2023.10.26.23297603

**Authors:** Tracey Smythe, Shanquan Chen, Sara Rotenberg, Marianne Unger, Emily Miner, Frederic Seghers, Chiara Servili, Hannah Kuper

## Abstract

**Background:** Play is essential for the cognitive, social, and emotional development of all children. Disparities potentially exist in access to play for children with disabilities, and the extent of this inequity is unknown.

**Methods:** Data from 212,194 children aged 2-4 years in 38 Low and Middle-Income Countries were collected in the UNICEF supported Multiple Indicator Cluster Survey (2017 – 2020). Disability was assessed by the Washington Group-Child Functioning Module. Logistic regression models were applied to investigate the relationship between disability and play opportunities, controlling for age, sex, and wealth status. Meta-analysis was used to pool the estimates (overall, and disaggregated by sex), with heterogeneity assessed by Cochran’s Q test.

**Findings:** Children with disabilities have approximately 9% fewer play opportunities than those without disabilities (adjusted RR [aRR] =0.88, 95%CI=0.82–0.93), and this varied across countries. Mongolia and Democratic Republic of São Tomé and Príncipe had the lowest likelihood of play opportunities for children with disabilities ((aRR=0.26,95%CI=0.09-0.75; aRR=0.46, 95%CI=0.23-0.93, respectively). Moreover, children with disabilities are 17% less likely to be provided with opportunities to play with their mothers (aRR=0.83, 95%CI: 0.73–0.93), which is further reduced for girls with disabilities (aRR=0.74, 95% CI:0.60-0.90) compared to their peers without disabilities. The associations varied by impairment type, and children with communication and learning impairments are less likely to have opportunities for play with aRR of 0.69 (95%CI: 0.60-0.79) and 0.78 (95%CI:0.71–0.86), compared to those without disabilities, respectively.

**Interpretation:** Children with disabilities are being left behind in their access to play and this is likely to have negative impacts on their overall development and well-being.

**Funding:** HK and TS receive funding from NIHR. The Study was funded by PENDA. SR receives funding from the Rhodes Trust.

**Evidence before this study:** We searched PubMed and Google Scholar for studies reporting population-representative estimates of children with disabilities’ exposure to play in low-income and middle-income countries (LMICs) published before Feb 7th, 2023. We used the following combination of keywords: “play” AND (“early childhood” OR “preschool” OR “young children”) AND (disability OR disabilities) AND “prevalence”. We found no multi-country studies reporting the prevalence or country-level disparities (within or between countries) for opportunities for play for children with disabilities. We did not identify any studies synthesising or comparing estimates across all dimensions of play or disability, nor did we identify studies reporting population-representative estimates of play for all LMICs. UNICEF has published global reports, which reveal that children with disabilities receive less early stimulation and responsive care and have limited exposure to children’s books and toys compared to children without disabilities, however opportunities for play within the household setting have not been examined in a comprehensive analysis.

**Added value of this study:** To our knowledge, this is the first study to explore the opportunities for play for children with disabilities compared to those without across multiple countries. Moreover, it provides a large dataset on this topic including 212,194 children aged 2–4 years from 38 low and middle income countries (LMICs), including approximately 6.1% with disabilities (i.e. reporting a lot of difficulty or more in a functional domain). The study has advanced the literature in five substantive ways. First, we highlighted that children with disabilities have fewer opportunities to play, across multiple measures of play and multiple settings. Second, we demonstrated that there are disparities in play opportunities for children with disabilities across countries. Third, we showed that this varied by impairment and was worst for children with learning and communication impairments. Fourth, we showed that there was a discrepancy between girls and boys with disabilities. Finally, our work extends beyond simple description by deploying ratio ratios to provide a quantitative risk assessment. This enables us to identify areas of particular concern and suggest where interventions may be most needed. The ratio ratios shed light on the severity of disparities and pinpoint specific high-risk categories such as particular countries, types of disabilities, or population groups. This analysis is crucial for refining interventions and optimising resource allocation, especially in low- and middle-income countries.

**Implications of all the available evidence:** The study findings emphasise the importance of including children with disabilities in early child development programmes, and where relevant preschool, which may require modifications to ensure inclusivity. Programmes are needed that specifically target children with learning or communication impairments. This may work best through parent support programmes, as formal preschool or programmes may be lacking in LMICs. Monitoring participation is crucial for children with disabilities. To promote equal opportunities for play at home, in schools, and in other community settings, it is necessary to improve the knowledge and attitudes of parents, teachers, and caregivers, as well as implement policies that address barriers to participation. The findings underscore the urgent need for policies to reflect the inclusion of children with disabilities. Research is needed to establish evidence regarding the importance of promoting play opportunities beyond the home environment, including pre-schools, schools, and community settings. Furthermore, well-designed studies to provide affordable, timely and accessible data on effective strategies for enhancing play for children with disabilities are required. This information will enable programme developers and policy makers to make evidence-based decisions on improving the lives of children with disabilities worldwide.

## Introduction

Play is a fundamental aspect of every child’s development and is essential for their cognitive, social, and emotional well-being and development ^1^. Ensuring that children have access to adequate play opportunities therefore is not only a moral obligation, but also has long-term benefits for children’s health and well-being, as well as their future success in education and employment ^2–4^. Nevertheless, not all children have access to, or experiences play in the same way, which can impair their development and well-being. Globally, certain groups, such as children with disabilities, may be particularly excluded from play due to factors such as limited resources, inadequate infrastructure, and societal attitudes towards disabilities ^5^. This challenge is shared by both low and middle-income countries, as well as high-income countries ^6–8^. For the purpose of this paper, play is defined as a voluntary, intrinsically motivated activity that involves active engagement, imagination, and exploration, providing opportunities for learning and social interaction ^9^. Disability refers to a broad range of impairments, that in interaction with personal and environmental factors, may affect a child’s physical, cognitive, sensory, or social functioning ^10^.

There are nearly 240 million children with disabilities globally, the majority of whom live in low and middle-income countries (LMICs). Available evidence suggests that many children with disabilities in LMICs face barriers that limit their opportunities to engage in play and early development activities ^11^. These barriers can be experienced at the individual, family or community level. At the individual level, challenges with communication and sensory processing can impact play and interaction with peers ^12^. Family level barriers include caregivers’ expectations and concerns about their child’s acceptance and interaction with peers, as well as limited self-efficacy in facilitating play for children with disabilities ^13^. Families and communities have an important role as caretakers for children with disabilities. Attitudinal barriers, such as negative attitudes and stereotypes towards disability that are held in the household, in the community, and at school, can limit expectations and opportunities for play ^14^. Lack of opportunities for interaction with peers and inadequate social support contribute to isolation and exclusion from social play experiences ^15, 16^. Furthermore, lack of appropriate play materials and limited access to outdoor spaces ^17^, as well as inaccessible play spaces and equipment that is not designed for their needs, can limit the ability of children with disabilities to engage in physical play and exploration ^18^. Together, the cumulative effect of these barriers can have a negative impact on the well-being and development of young children with disabilities, making it essential to prioritise play opportunities and foster inclusive environments that cater to their specific needs and abilities ^19^.

There is pressing need to increase attention to the barriers that hinder play and recognise the benefits it brings in fostering the development of children with disabilities, particularly in early childhood. Opportunities for play are important aspects of children’s participation ^20^ that can also contribute to their quality of life, functional independence and life satisfaction ^20–22^. The recent inclusion of the Washington Group/UNICEF Child Functioning Module in the Multiple Indicator Cluster Survey (MICS) ^23^ has produced some of the first standardised and internationally comparable nationally representative estimates of child disability and child functioning in many LMICs. A recent UNICEF report (2021) ^16^ combined these data to document inequities in ‘early stimulation and responsive care’ concerning disability. Building on this analysis, we explore the unequal distribution of opportunities to play within the household setting among girls and boys, categorised by disability, in each country.

It is crucial to understand the opportunities and barriers faced by children with disabilities in order to promote inclusive and accessible opportunities for play. This understanding becomes even more imperative when considering the intersection of gender and disability, especially for young children ^24^. The unique experiences and challenges faced by girls and boys with disabilities can have an impact on their ability to fully participate in play activities and receive necessary support ^25–27^. Moreover, there also needs to be a particular focus on LMICs as data is lacking ^28–31^. The focus on LMICs is crucial as 80% of people with disabilities reside in these countries. The level of exclusion, such as from schools, is likely to be particularly high in these settings, where health systems are also weaker^10^. Therefore, emphasising what happens within the home environment becomes particularly important. The objective of this study is to examine caregivers’ engagement in various play activities with their children aged 2 to 4. We aim to investigate the opportunities for play among young children with disabilities compared to young children without disabilities in LMICs.

## Methods

### Data source

The dataset used in this study is based on the UNICEF-supported MICS, a comprehensive survey programme conducted in LMICs to collect nationally and sub-nationally representative data on key health and development indicators ^32^. The MICS employs a cross-sectional, household survey approach conducted by national statistical authorities with support from UNICEF. The surveys used standardised questionnaires administered by trained data collectors who interview mothers or primary caregivers about their children. The MICS is a crucial data source for monitoring progress on the Sustainable Development Goals (SDGs), employing a multi-stage sampling methodology to generate representative estimates at the national, regional, and urban-rural levels ^33^. For this study, individual participant data from the sixth round of the MICS (conducted 2017-2020) were utilised. This round incorporated the Washington Group Questions, enabling the disaggregation of data based on disability status. We selected MICS surveys which included data on our variables of interest and which were publicly available on the UNICEF website in March 2023, which included 47 LMICs (covering 289,616 children aged in range 2–4).

### Participants

Children with disabilities, aged two to four, are identified using the Child Functioning Module (CFM), a module developed and validated by UNICEF and the Washington Group on Disability Statistics ^34^. Caregivers respond to questions regarding their child’s difficulties across eight domains (seeing, hearing, walking, fine motor skills, communication, learning, playing, and controlling behaviour). Caregivers respond to each question and assess the level of difficulty their child experiences from ‘no difficulty’, ‘some difficulty, ‘a lot of difficulty’ or ‘cannot do at all’. Those children for whom caregivers reported ‘cannot do at all’ or have ‘a lot of difficulty’ (or ‘a lot more’ with regard to the variable ‘controlling behaviour’) in at least one domain were considered to be children with disabilities.

Missing values for the studies variables (details as following) ranged from 5.7% (for play overall [at least 4 times]) to 14.8% (for disability). We omitted the individuals with missing values instead of any imputation.

To reduce the bias due to the small sample size, following the suggestion of UNICEF, we excluded countries with fewer than 25 respondents with disabilities when we did the country-specific estimations. Appendix Figure 1 shows the flowchart of selection of participants in detail.

**Figure 1:**
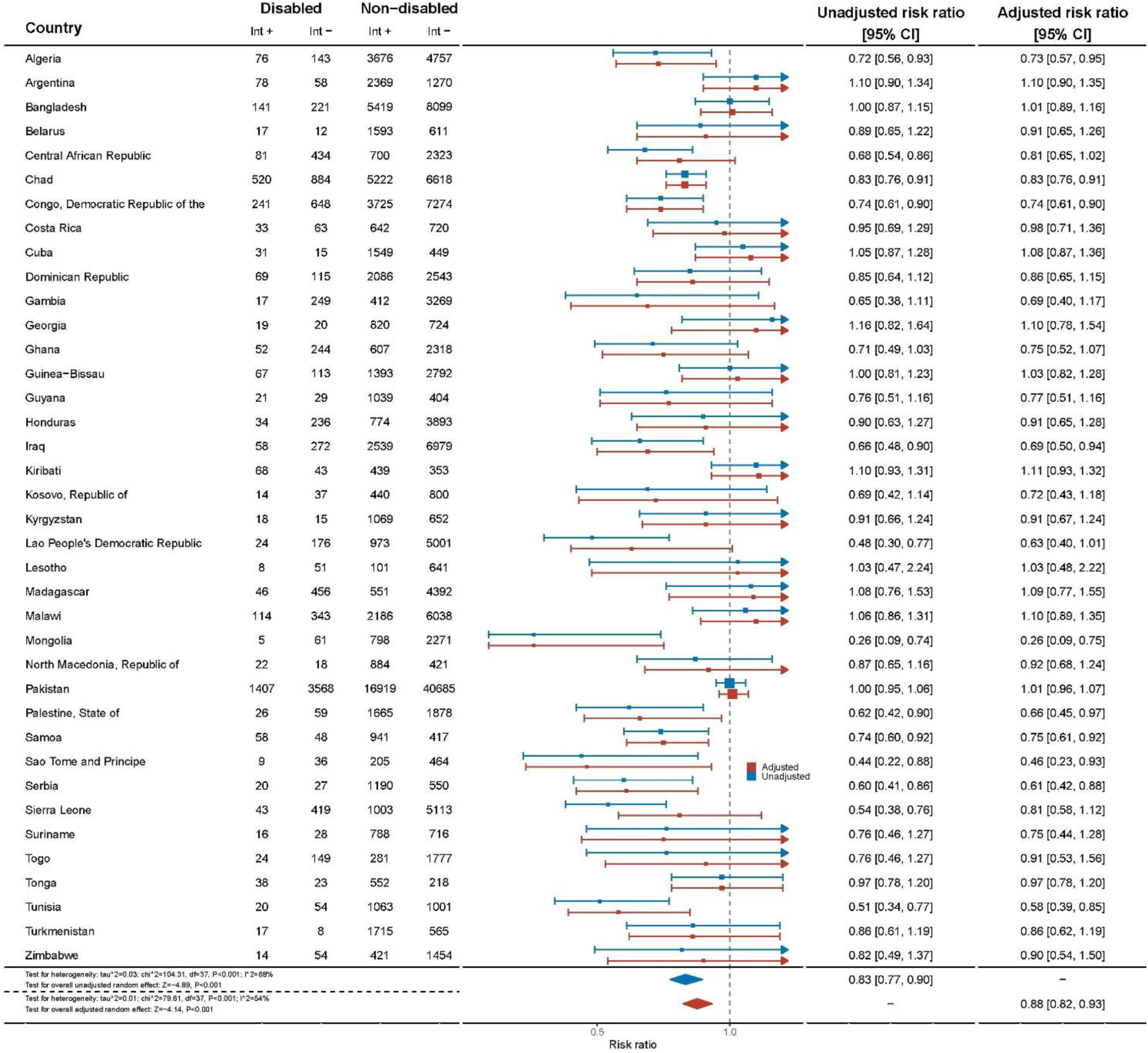
Play opportunities by country*. *Data were extracted from weighted logistic regression models with ‘opportunity to play’ as the outcome and disability status as the key predictor, controlling for age, sex, and wealth status.

### Outcome of interest

The opportunity for play was assessed using four MICS-defined variables that included questions focussed on: telling stories; singing songs; taking the child outside the home; and playing with the child. In this study, offering all four of these activities within the last three days was considered as providing an opportunity to play (Appendix 2). This definition of ‘opportunities to play’ is a subset of six variables that fall under the broader umbrella of “early stimulation and responsive care” as defined by UNICEF ^16^. The early stimulation and responsive care indicator’s definition is the “percentage of children aged 24 to 59 months who engaged in four or more activities to provide early stimulation and responsive care in the last three days with any adult household member. Activities include reading books or looking at picture books with the child; telling stories; singing songs to or with the child; taking the child outside the home; playing with the child; naming, counting or drawing things for or with the child ^16^.”

### Covariates

In this study, we have considered following socio-demographic factors, given their potential association with opportunities to play, including age (years), sex, living with parents (both, only mother, only father, or none), number of children, education level of mother and father (pre-primary or none, primary, secondary, or higher secondary), wealth status (top 40% vs not), and residence place (urban vs rural). Wealth indices were constructed using household characteristics, including ownerships of goods, living situation, water and sanitation, and other assets^35^. Number of children was collected from the responding caregiver by two questions on the number of children under age 5 and number of children age 5-17.

### Data analysis

The continuous data in the study were reported as mean values and standard deviations (SD), while the between-group difference was assessed using the t-test. Categorical data was presented in terms of the number of cases and percentages. The between-group difference for the categorical data was evaluated using the chi-square test.

In order to investigate the relationship between disability and opportunities to play, a weighted logistic regression model was applied for each country. The dependent variable was ‘opportunity to play’, which was classified as either “yes” or “no”. Disability status, also categorized as “yes” or “no” with “no” as the reference, was considered the key predictor. The models were adjusted for child age, child sex, and family wealth status (being in the top 40% wealth bracket or not). We accounted for survey design and sample weights provided by UNICEF. The unadjusted and adjusted results were presented as risk ratios (RR) with their corresponding 95% confidence intervals (CI). Subsequently, a meta-analysis was performed to pool the estimates from all countries, with the inverted standard error serving as the weight. Cochran’s Q test was employed to assess the heterogeneity of estimates across countries ^36^. If significant heterogeneity (p < 0.1) was present, a random-effect meta-analysis was conducted; otherwise, a fixed-effects meta-analysis was employed. The logistic regression models were repeated, adding an interaction term of sex and disability to examine whether the association between disability and opportunity to play was moderated by sex.

Similar analyses were carried out to investigate the relationship between subtypes of disability and opportunity to play, and opportunities to play with specific individuals, such as mothers, fathers, and other caregivers, using the same weighted logistic regression models.

We conducted a sensitivity analysis. To account for individuals with missing values, we conducted 20 multiple imputations using chained equations and repeated our analyses. The analyses were performed using R version 4.2.2, and statistical significance was determined as p < 0.05.

## Results

A total of 212,194 children from 38 countries between the ages of 2 and 4 years were included in the study. This sample included approximately 12,995 (6.1%) children with disabilities (**Table 1**). Forty-five percent (n=5,824) of children with a disability were girls. The most common type of reported difficulties was with behaviour, affecting 4,654 children with disabilities (35.8%), followed by learning (n=4,002, 30.8%) and communication (n=3,875, 29.8%). Hearing was the least common type of difficulty affecting approximately 6.5% (n=839) of children with disabilities. There are sex differences in specific difficulties among children with disabilities, with higher levels of reported learning (p<0.001) and hearing (p=0.034) difficulties among girls with disabilities, and higher levels of reported behavioural difficulties (p<0.001) among boys with disabilities. Socio-economic status was poorer for families of children with disabilities (p<0.001), as measured by maternal education or wealth status.

**Table 1:** Characteristics of children aged 2-4years and their families*.

| Characteristic | All children in sample (N =212194) |  |  | All children with disabilities (n=12995) |  |  |
| --- | --- | --- | --- | --- | --- | --- |
|  | No disability | Disability | P | Girls with disability | Boys with disability | P |
|  | (n = 199199) | (n = 12995) |  | (n=5824) | (n=7171) |  |
| **Age (years)** | 3.02 (0.81) | 2.87 (0.82) | <0.001 | 2.86 (0.82) | 2.88 (0.82) | 0.167 |
| **Sex (= girls)** | 97838 (49.1) | 5824 (44.8) | <0.001 | - | - | - |
| **Type of disability<sup>^</sup>** |  |  |  |  |  |  |
| Behaviour | - | 4654 (35.8) | - | 1929 (33.1) | 2725 (38.0) | <0.001 |
| Learning | - | 4002 (30.8) | - | 1898 (32.6) | 2104 (29.3) | <0.001 |
| Communication | - | 3875 (29.8) | - | 1752 (30.1) | 2123 (29.6) | 0.567 |
| Walking | - | 1765 (13.6) | - | 806 (13.8) | 959 (13.4) | 0.456 |
| Playing | - | 1585 (12.2) | - | 719 (12.3) | 866 (12.1) | 0.661 |
| Seeing | - | 1465 (11.3) | - | 672 (11.5) | 793 (11.1) | 0.405 |
| Fine motor | - | 1172 (9.0) | - | 532 (9.1) | 640 (8.9) | 0.701 |
| Hearing | - | 839 (6.5) | - | 406 (7.0) | 433 (6.0) | 0.034 |
| **Living with parents** |  |  |  |  |  |  |
| Both | 173830 (87.3) | 11300 (87.0) | 0.648 | 5042 (86.6) | 6258 (87.3) | 0.673 |
| Only mother | 18399 (9.2) | 1223 (9.4) |  | 563 (9.7) | 660 (9.2) |  |
| Only father | 2562 (1.3) | 166 (1.3) |  | 75 (1.3) | 91 (1.3) |  |
| None | 4408 (2.2) | 306 (2.4) |  | 144 (2.5) | 162 (2.3) |  |
| **Number of children** | 3.98 (2.75) | 4.51 (3.23) | <0.001 | 4.57 (3.28) | 4.47 (3.19) | 0.077 |
| **Education level of Mother** |  |  |  |  |  |  |
| Pre-primary or none | 73687 (37.0) | 6489 (49.9) | <0.001 | 3037 (52.1) | 3452 (48.1) | <0.001 |
| Primary | 47659 (23.9) | 3024 (23.3) |  | 1300 (22.3) | 1724 (24.0) |  |
| Secondary | 36480 (18.3) | 1820 (14.0) |  | 785 (13.5) | 1035 (14.4) |  |
| Higher secondary | 41373 (20.8) | 1662 (12.8) |  | 702 (12.1) | 960 (13.4) |  |
| **Education level of Father** |  |  |  |  |  |  |
| Pre-primary or none | 45586 (22.9) | 4153 (32.0) | <0.001 | 1962 (33.7) | 2191 (30.6) | 0.002 |
| Primary | 42347 (21.3) | 2676 (20.6) |  | 1168 (20.1) | 1508 (21.0) |  |
| Secondary | 37094 (18.6) | 2016 (15.5) |  | 877 (15.1) | 1139 (15.9) |  |
| Higher secondary | 74172 (37.2) | 4150 (31.9) |  | 1817 (31.2) | 2333 (32.5) |  |
| **Wealth status (= top 40%)** | 61825 (31.0) | 3385 (26.0) | <0.001 | 1448 (24.9) | 1937 (27.0) | 0.006 |
| **Residence place (= urban)** | 65690 (33.0) | 3448 (26.5) | <0.001 | 1538 (26.4) | 1910 (26.6) | 0.786 |

\* Data are presented as mean (sd) for continuous variables, tested by t-test; and as number (percentage) for categorical variables, tested by chi-square test.
<sup>^</sup> the percentages do not add up to 100% as children could have multiple disabilities or present with overlapping difficulties in the text, and in a footnote

Children with disabilities are less likely to have opportunities to play compared to children without disabilities (27.4% vs 34.5%, p<0.001). Girls with disabilities are also less likely to have play opportunities compared to boys with disabilities (p=0.031) (**Table 2**). Furthermore, children with disabilities overall experience fewer play opportunities to play with their mother (10.8% vs 16.7%, p<0.001) and father (6.0% vs 7.7%, p<0.001)) in comparison to their counterparts without disabilities. Girls with disabilities have fewer opportunities to play with their mother (p=0.028) and father (p=0.008) compared to boys with disabilities.

**Table 2:** Characteristics of play opportunities for children aged 2-4 years.

| Characteristic | Children without disability N (%) | Children with disability N(%) | P | Girls with disabilities | Boys with disabilities | P |
| --- | --- | --- | --- | --- | --- | --- |
| **Opportunity for play, number (%)** | 68749 (34.5) | 3566 (27.4) | <0.001 | 1543 (26.5) | 2023 (28.2) | 0.031 |
| **Play with specific individuals** |  |  |  |  |  |  |
| Play opportunities with mother | 33334 (16.7) | 1397 (10.8) | <0.001 | 587 (10.1) | 810 (11.3) | 0.028 |
| Play opportunities with father | 10157 (5.1) | 498 (3.8) | <0.001 | 194 (3.3) | 304 (4.2) | 0.008 |
| Play opportunities with other people | 15333 (7.7) | 782 (6.0) | <0.001 | 339 (5.8) | 443 (6.2) | 0.416 |

Adjusted analyses show that on average, children with disabilities had approximately 9% fewer play opportunities than those without disabilities (adjusted RR[aRR]=0.88, 95%CI=0.82–0.93). This difference is consistent for both girls with disabilities (aRR=0.86, 95%CI=0.78-0.94) and boys with disabilities (aRR=0.90, 95%CI=0.82-0.98) compared to their peers without disabilities. Children with disabilities are 17% less likely to have play opportunities with their mother (aRR 0.83, 95%CI: 0.73–0.93) compared to those without disabilities, and for girls with disabilities the likelihood is even lower (aRR 0.74, 95%CI: 0.60-0.90). Children with disabilities also face reduced likelihoods of reduced play opportunities with other people (aRR0.86, 95%CI: 0.78-0.93) and this is similar for both girls and boys with disabilities when compared to their counterparts (**Table 3**). Girls with disabilities have increased play opportunities with their fathers when compared to boys with disabilities (aRR 7.80, 95%CI: 1.14-53.36) (**Appendix 3**).

**Table 3:** Adjusted risk ratio for play opportunities experienced by children (2-4 years) with disabilities*.

| Outcomes | Overall aRR | Girls with disabilities aRR | Boys with disabilities aRR |
| --- | --- | --- | --- |
| **Opportunity for play** | **0.88 [0.82, 0.93] &** | **0.86 [0.78, 0.94] &** | **0.90 [0.82, 0.98] &** |
| **Play with specific individuals** |  |  |  |
| Play opportunities with mother | **0.83 [0.73, 0.93] &** | **0.74 [0.60, 0.90] &** | 0.87 [0.75, 1.01] |
| Play opportunities with father | 0.80 [0.64, 1.01] | 0.77 [0.55, 1.10] | 0.82 [0.62, 1.08] |
| Play opportunities with other people | **0.86 [0.78, 0.93] &** | **0.87 [0.76, 0.99] &** | **0.84 [0.74, 0.94] &** |

\* Data presented as the adjusted Risk Ratio(aRR) and its 95% confidence interval (CI), extracted from meta-analysis of country-specific results from weighted logistic regression, which took variable in column as outcome, disability as the key predictor, controlling for child age, child sex, and family wealth status.
& $p < 0.05$

Play opportunities for children with disabilities vary across countries (**Fig 1**). Children with disabilities in Mongolia and Democratic Republic of São Tomé and Príncipe had the lowest likelihood of play opportunities (aRR=0.26,95%CI=0.09-0.75; aRR=0.46, 95%CI=0.23-0.93, respectively), whilst children with disabilities in Kiribati had the highest likelihood (aRR=1.11, 95%CI=0.93-1.32) (**Fig 1**). There is moderate heterogeneity among the data, with an I^2 statistic of 68% and a test for heterogeneity p-value of less than 0.001.

There are notable disparities in the provision of play opportunities by mothers to children with disabilities across various countries. In Guinea-Bissau, children with disabilities had a 66% lower likelihood (aRR=0.34, 95%CI=0.18-0.65) of receiving play opportunities from their mothers compared to children without disabilities. Similarly, in Democratic Republic of Congo and in Ghana, children with disabilities had over 50% fewer play opportunities (aRR=0.45, 95%CI=0.31-0.66; aRR=0.43, 95%CI=0.17-1.06, respectively) with their mother than their non-disabled peers. In contrast, in Pakistan, children with disabilities were 31% more likely to receive play opportunities from their mothers compared to those without disabilities (aRR=1.31, 95%CI=1.18-1.45) (**Fig 2**).

**Figure 2:**
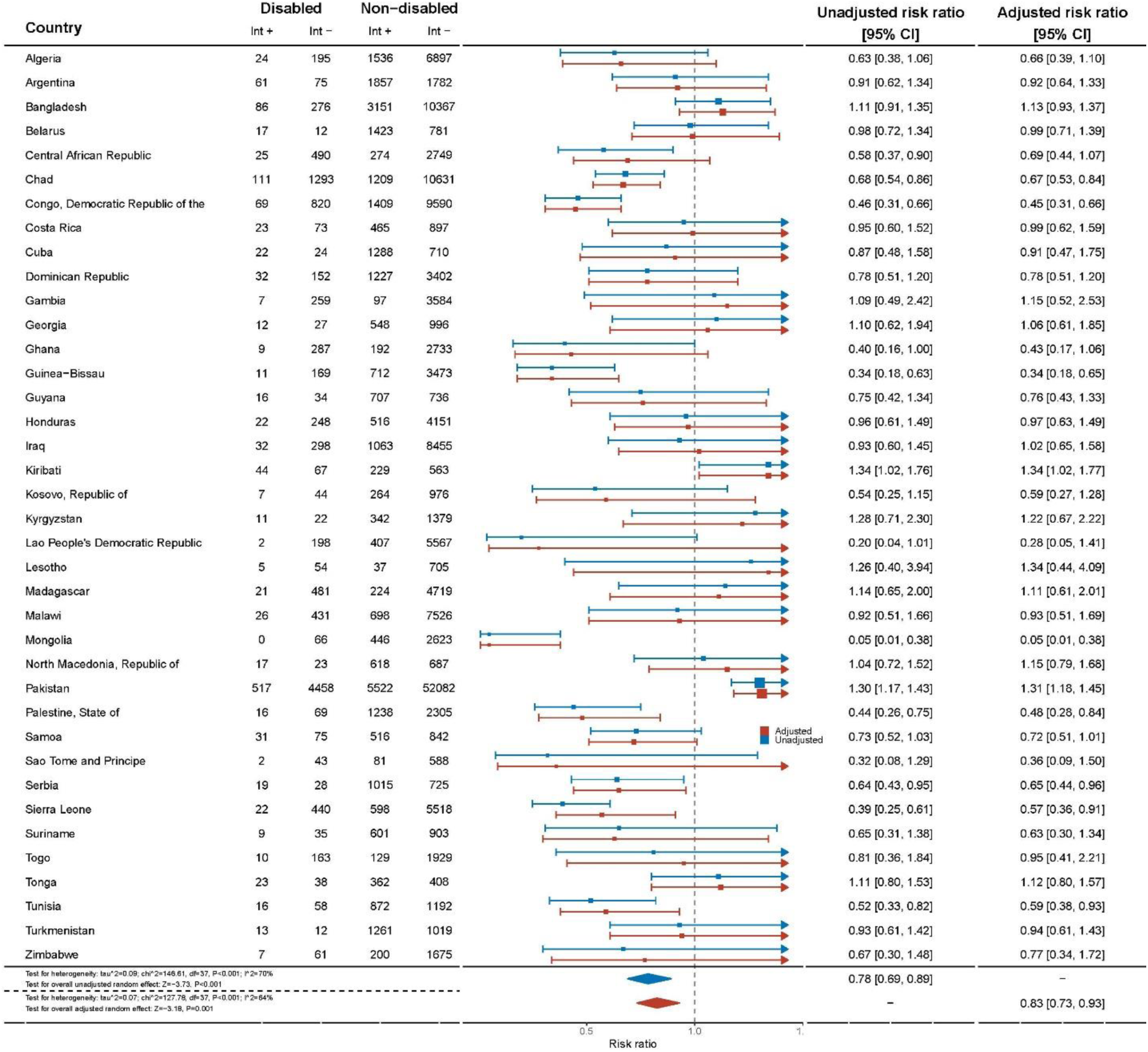
Opportunity to play provided by mother disaggregated by country.

The variation in play opportunities provided by fathers also exhibited significant differences across countries. In Cuba, children with disabilities had a significantly lower likelihood of receiving play opportunities from their fathers, with an adjusted risk ratio of 0.21 (95%CI=0.07–0.61), compared to children without disabilities. In Serbia, children with disabilities faced a similar challenge, with an aRR of 0.31 (95%CI=0.11-0.84), compared to their counterparts without disabilities. On the other hand, children in Pakistan and Kiribati experienced more favourable outcomes, as they had a higher likelihood of receiving play opportunities from their fathers (Pakistan: aRR=1.50, 95%CI=1.28-1.75; Kiribati: aRR=1.78, 95%CI=1.21-2.62), compared to children without disabilities (**Fig 3**).

**Figure 3:**
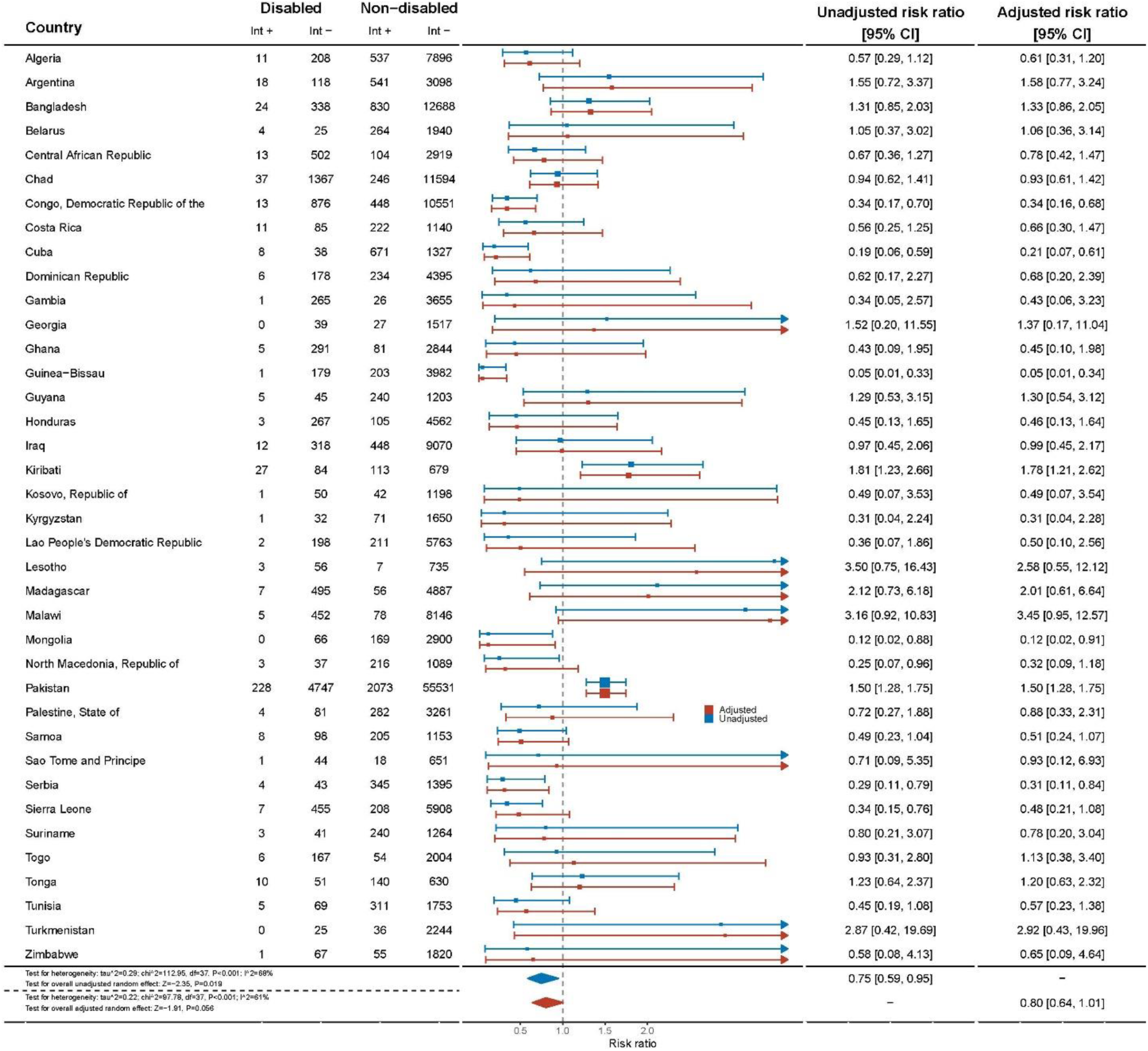
Opportunity to play provided by father disaggregated by country.

Similarly, children with disabilities had a 14% reduced likelihood of being provided with opportunities to play by other people compared to children without disabilities (aRR=0.86, 95%CI=0.78-0.93) and this varied by country (**Appendix 4**).

Children with communication difficulties (aRR=0.69, 95%CI=0.60-0.79) and learning difficulties (aRR=0.78, 95%CI=0.71–0.86) had reduced likelihood of opportunities for play compared to those without disabilities, and this was also reduced for play opportunities provided by their mothers (aRR=0.68, 95%CI:0.57-0.81; aRR=0.65, 95%CI: 0.53-0.80 respectively) (**Table 4**). Children with other types of difficulties had no difference in opportunities for play compared to their peers without disabilities. However, there were increased play opportunities offered to children with visual impairment by mothers (aRR=1.46, 95%CI=1.10-1.93), fathers (aRR=2.33, 95%CI=1.55-3.50) and other people (aRR=2.40, 95%CI=1.47-3.94). Children with hearing impairment also experienced an increase in play opportunities compared to those without disabilities when mothers (aRR=1.43, 95%CI=1.10-1.86) and fathers (aRR=3.93, 95%CI=2.75-5.61) were involved, with a similar increase provided by other people (aRR=3.23, 95%CI=2.14-4.87). Children with walking, fine motor and play impairments experienced an increase in opportunities to play provided by fathers and other people.

**Table 4:** Play experiences disaggregated by impairment type (with children without disabilities as reference)*

| Impairment | Opportunity to play overall [95%CI] | Mother provides play [95%CI] | Father provides play [95%CI] | Other people provide play [95%CI] |
| --- | --- | --- | --- | --- |
| Seeing | 1.17 [0.96, 1.42] | **1.46 [1.10, 1.93] &** | **2.33 [1.55, 3.50] &** | **2.40 [1.47, 3.94] &** |
| Hearing | 1.07 [0.92, 1.24] | **1.43 [1.10, 1.86] &** | **3.93 [2.75, 5.61] &** | **3.23 [2.14, 4.87] &** |
| Walking | 0.96 [0.79, 1.16] | 1.21 [0.93, 1.57] | **1.98 [1.34, 2.92] &** | **2.14 [1.51, 3.04] &** |
| Fine motor | 0.93 [0.71, 1.21] | 1.37 [0.96, 1.96] | **3.50 [2.25, 5.43] &** | **2.76 [1.70, 4.45] &** |
| Communication | **0.69 [0.60, 0.79] &** | **0.68 [0.57, 0.81] &** | 0.77 [0.58, 1.02] | 0.88 [0.66, 1.16] |
| Learning | **0.73 [0.65, 0.82] &** | **0.65 [0.53, 0.80] &** | 0.77 [0.54, 1.10] | 0.82 [0.63, 1.06] |
| Play | 0.87 [0.75, 1.01] | 1.05 [0.85, 1.29] | **1.79 [1.22, 2.64] &** | **1.71 [1.15, 2.54] &** |
| Behaviour | 1.04 [0.95, 1.13] | 0.93 [0.80, 1.08] | 1.23 [0.89, 1.68] | 1.16 [0.92, 1.45] |

\* Data presented as the Risk Ratio(RR) and its 95% confidence interval (CI), extracted from meta-analysis of country-specific results from weighted logistic regression, which took variable in column as outcome, disability as the key predictor, controlling for child age, child sex, and family wealth status.
& p < 0.05

The sensitivity analysis on missing values with multiple imputations by chained equations confirmed the consistence of the above results (**Appendix 5**).

## Discussion

This study examined play opportunities for 212,194 children aged between 2-4 years from 38 LMICs, including 6.1% with disabilities. We found that children with disabilities were less likely to have opportunity to play compared to those without disabilities, and this difference was consistent for both girls and boys with disabilities. The study also revealed that children with disabilities, in particular girls, were less likely to have play opportunities offered by their mother. Both girls and boys with disabilities equally experience fewer opportunities to play with other people. Furthermore, children with communication and learning impairments were less likely to have opportunities for play compared to their peers without disabilities.

However, children with visual and hearing impairments experienced increased opportunities to play compared to their counterparts without disabilities, particularly when mothers and fathers were involved. Children with walking, fine motor, and play impairments encountered enhanced play opportunities with fathers and other people.

The study also revealed global variations, with certain countries such as Mongolia and Democratic Republic of São Tomé and Príncipe, offering lower play opportunities, while others, like Kiribati, showed more favourable opportunities for children with disabilities to engage in play. Pakistan, when analysed based on involvement by the mother, father or other individuals, displayed a higher likelihood of play opportunities for children with disabilities.

These variations highlight the importance of considering local contexts and implementing targeted interventions based on the specific needs and challenges faced by children with disabilities in different regions. Moreover, the variations across countries suggest that cultural, societal, and systemic factors may contribute to the uneven distribution of play opportunities for children with disabilities. For example, with regard cultural and societal factors, varying levels of awareness, acceptance, and inclusivity towards children with disabilities in different societies can influence the provision of play opportunities. Societal attitudes and norms of disability may impact the allocation of resources ^37^ towards supporting inclusive play initiatives, such as the establishment of community-based programs and parent support groups. Countries that prioritise disability rights, inclusive policies, and community engagement may be more likely to offer enhanced play opportunities for children with disabilities ^38^. Systemic influences that also contribute to these disparities may include variations in healthcare systems, social welfare programs, and educational policies across countries, which can affect the availability of early intervention services that facilitate play for children with disabilities ^37^.

Whilst opportunities to play in early childhood, especially with parental engagement and support, promotes motor development ^39^, cultural expectations, traditional gender roles, and social norms can shape attitudes and involvement in play. Parents may encounter barriers or lack awareness regarding the importance of play for children with disabilities ^40^. Additionally, structural factors, such as work demands, can impact the extent of parents’ involvement in play ^41–43^. Children with disabilities, both girls and boys, faced a notable disparity in play opportunities offered by other people. These complex dynamics necessitate further exploration to understand the underlying reasons (for example, stigma ^43–45^) and to develop strategies that promote active engagement in play activities for children with disabilities. For instance, in Pakistan, mothers and fathers actively engaged in providing play opportunities for children with disabilities, while in Kiribati other people offered more play opportunities to children with disabilities compared to their peers without. Other countries could consider examining the models of engagement offered in Pakistan and Kiribati, taking a culturally sensitive approach, to foster more supportive play opportunities.

Additionally, children with communication and learning impairments face even greater limitations in accessing play opportunities compared to those without these impairments. Communication and learning impairments can create additional barriers to participating in play activities that rely on verbal communication or cognitive skills. Consequently, children with these impairments may require specialized strategies and resources to fully engage in play. For instance, visual supports such as communication boards and simple adaptations to games, such as using tactile materials or incorporating sensory elements, can enhance the involvement of children with communication and learning impairments. These examples highlight the importance of targeted interventions and support for this specific subgroup.

The absence of a significant difference between girls and boys with disabilities in terms of their play opportunities overall is a noteworthy finding. Despite the intersecting marginalisation typically faced by girls with disabilities^46^, this study suggests that both genders experience similar disparities in play opportunities overall. Nevertheless, there were disparities between boys and girls when offered play opportunities by their mother, with less opportunities being offered to girls. This underscores the importance of considering the unique obstacles encountered by all children with disabilities, regardless of their gender, to ensure equitable access to play and other essential opportunities.

The findings of our study contribute to the existing literature on the limited play opportunities available to children with disabilities, particularly in LMIC. While play is known to have positive impacts on physical, social, and cognitive development, our results highlight that children with disabilities may have limited opportunities for play compared to their peers without disabilities. This is consistent with previous research that has documented barriers to play for children with disabilities, such as a lack of resources such as books or play materials, as well as negative attitudes towards disability and lack of knowledge ^6, 12, 17, 47, 48^. The identified variation in play opportunities across countries could support this explanation to some extent. In addition, our study shows the specific challenges faced by children with communication and learning impairments in accessing play activities. This is an important finding as children with these types of impairments may require specialised support to fully engage in play. Overall, our study emphasises the need for increased attention and resources to address the play needs of children with disabilities in LMIC. Efforts to promote inclusive and accessible play opportunities, as well as education for caregivers on the importance of play for children’s development, can help to narrow the gap in play experiences between children with and without disabilities in these settings ^49^. Such interventions may ultimately enhance the well-being and quality of life of children with disabilities in LMIC ^50, 51^. Promoting the inclusion of children with disabilities in preschools may provide further opportunities for play, in settings where these are available.

This study has implications for policy makers, and opportunities exist to expand planning for inclusive early development through WHO guidelines and frameworks, which shape governments’ health expenditure, development assistance and targets. For example, in the adaptation of WHO’s Caregiver skills training for families of children with developmental delays and disabilities ^52, 53^ and the development of other elements of the caregiver skills training programme present avenues for action. For instance, based on our findings, the adaptation of the WHO Caregiver Skills Training could include specific modules that address the unique play needs of children with disabilities. These modules could focus on promoting play opportunities through targeted strategies, such as providing adaptive play materials, incorporating sensory stimulation, and encouraging peer interaction. By integrating these evidence-based recommendations into the existing framework, caregivers can be equipped with the knowledge and skills necessary to foster inclusive play environments that support the development and well-being of children with disabilities. Collaboration and effective leadership are crucial for supporting UNICEF with the monitoring of the developmental progress of children under five years of age in terms of health, learning, and psychosocial well-being (SDG 4.2.1) ^29^. This is necessary to achieve optimal early childhood development for children with disabilities, in line with the goal of ensuring that all children have access to quality early childhood development, care, and pre-primary education by 2030 (SDG 4.2)^54^. Play is a critical component of the development and well-being of children with disabilities, particularly for young children aged 2-4 years ^55–57^.

This study has limitations to consider when interpreting the results. Firstly, there was no data provided for children under 2 years, even though the first 24 months of development are particularly critical for stimulation. The study did not examine what “reduced opportunities for play” meant in terms of actual access to play for children with disabilities, especially since the results were reported by parents without any quality measure (i.e., time spent, frequency, appropriateness of content for age, etc.). As such, further research is necessary to understand the drivers of disparities and effective interventions. Furthermore, the instrument, CFM, uses umbrella terms to collect information on disabilities, and lacks specific examples of its subtypes. (e.g., type of behavioural difficulties). Children with milder levels of disability may be less likely to be identified by their caregivers, although they may also benefit from play. There were also important strengths, including the use of a large and multi-country dataset, detailed and standardised measures of disability and play, and the emphasis on data from LMICs. Finally, we acknowledge that although our study design and analytical approach were aimed at robustly assessing the associations of interest, the risk of type I errors due to multiple comparisons remains a limitation.

## Conclusion

Children with disabilities experience disparities in play opportunities compared to their peers without disabilities. Children with communication and learning impairments face particularly severe limitations in accessing play opportunities. These disparities are an affront to the rights of children with disabilities and will likely negatively impact their development and wellbeing. There is an urgent need to address social, attitudinal, and support barriers to ensure that children with disabilities can fully experience the benefits of play and thrive. Efforts should be made to promote inclusive play environments, provide access to adapted play resources, and raise awareness about the importance of play for all children, regardless of their abilities. This study contributes by specifically investigating play opportunities within the household setting, highlighting the urgency of addressing these disparities and advocating for inclusive play experiences for children with disabilities.

## Data Availability

All data produced in the present study are available upon reasonable request to the authors

## Appendix 1

**Figure 1:**
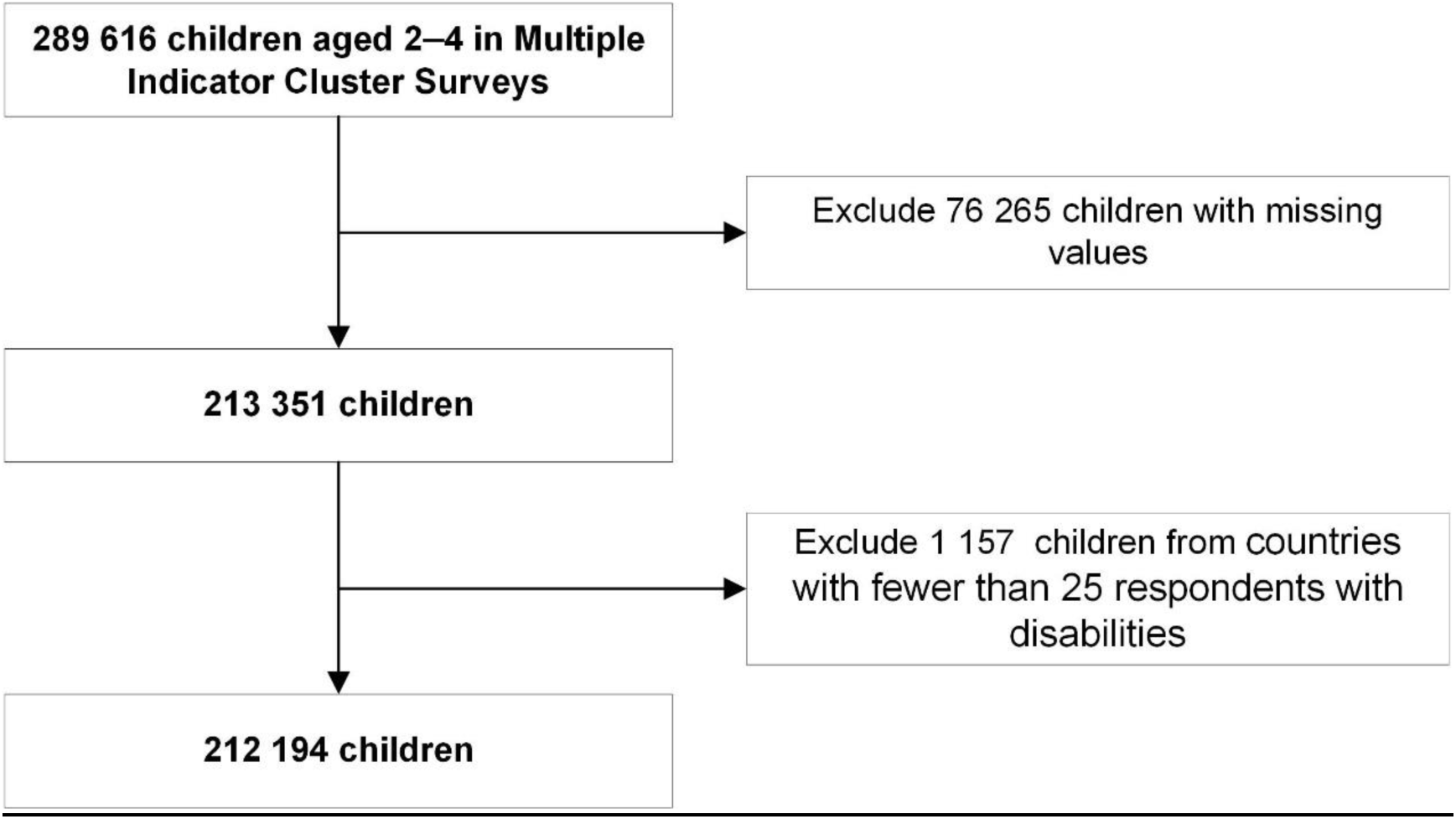
flowchart of selection of participants

## Appendix 2: Opportunities for play definition

Engagement of four or more activities in the last three days: Activities include reading books or looking at picture books with the child; telling stories; singing songs to or with the child; taking the child outside the home; playing with the child; naming, counting or drawing things for or with the child.

Example question:

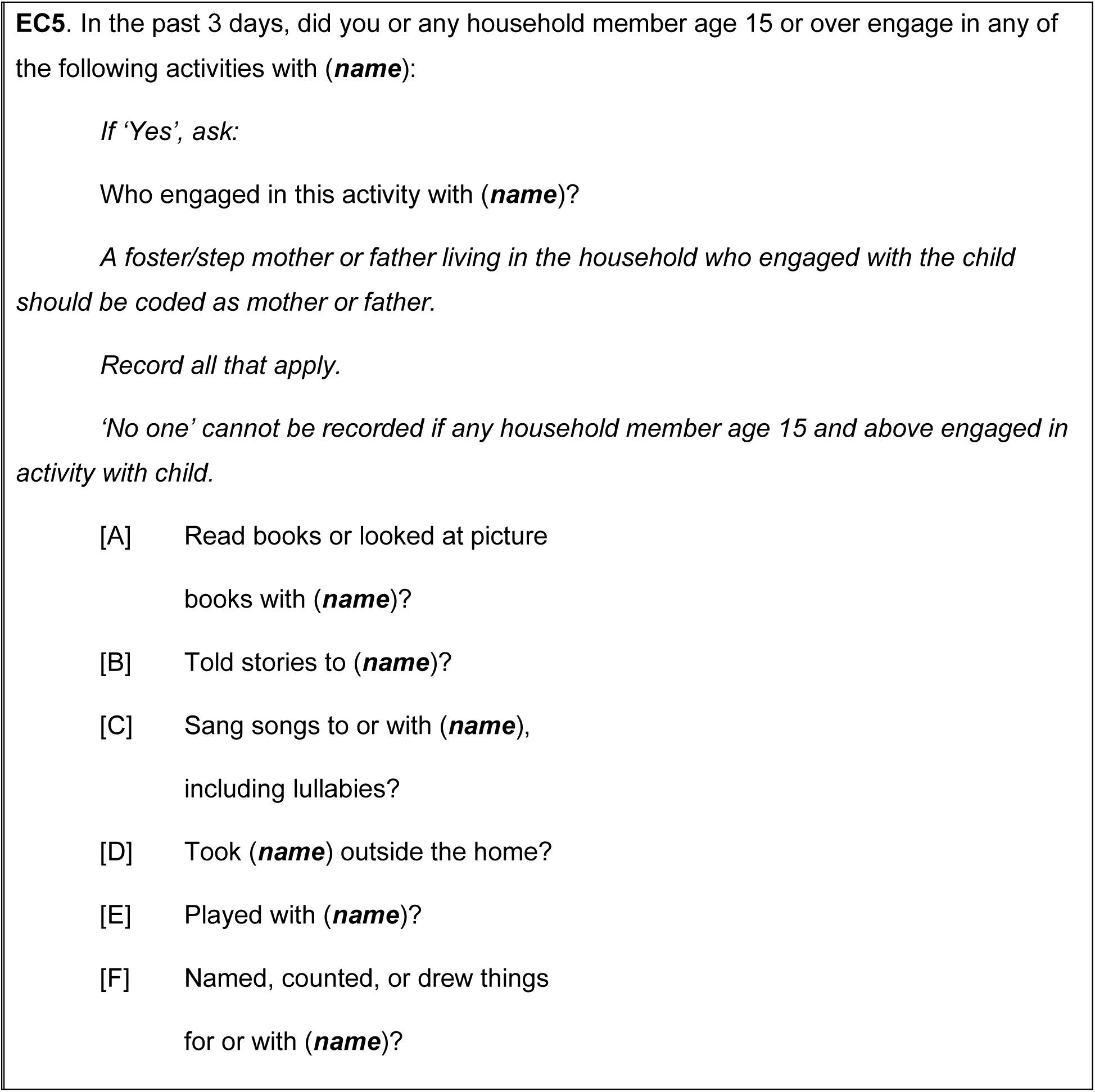

## Appendix 3: Play opportunities for boys and girls with disabilities

| Variable | Play Opportunities | Play opportunities with mother | Play opportunities with father | Play opportunities with other people |
| --- | --- | --- | --- | --- |
| Boys with disability | Reference | Reference | Reference | Reference |
| Girls with disability | 0.97 [0.90, 1.03] | 0.90 [0.78, 1.03] | **7.80 [1.14, 53.36]**<br>& | 1.92 [0.83, 4.44] |

& p < 0.05

## Appendix 4: Opportunities for play with other people disaggregated by country

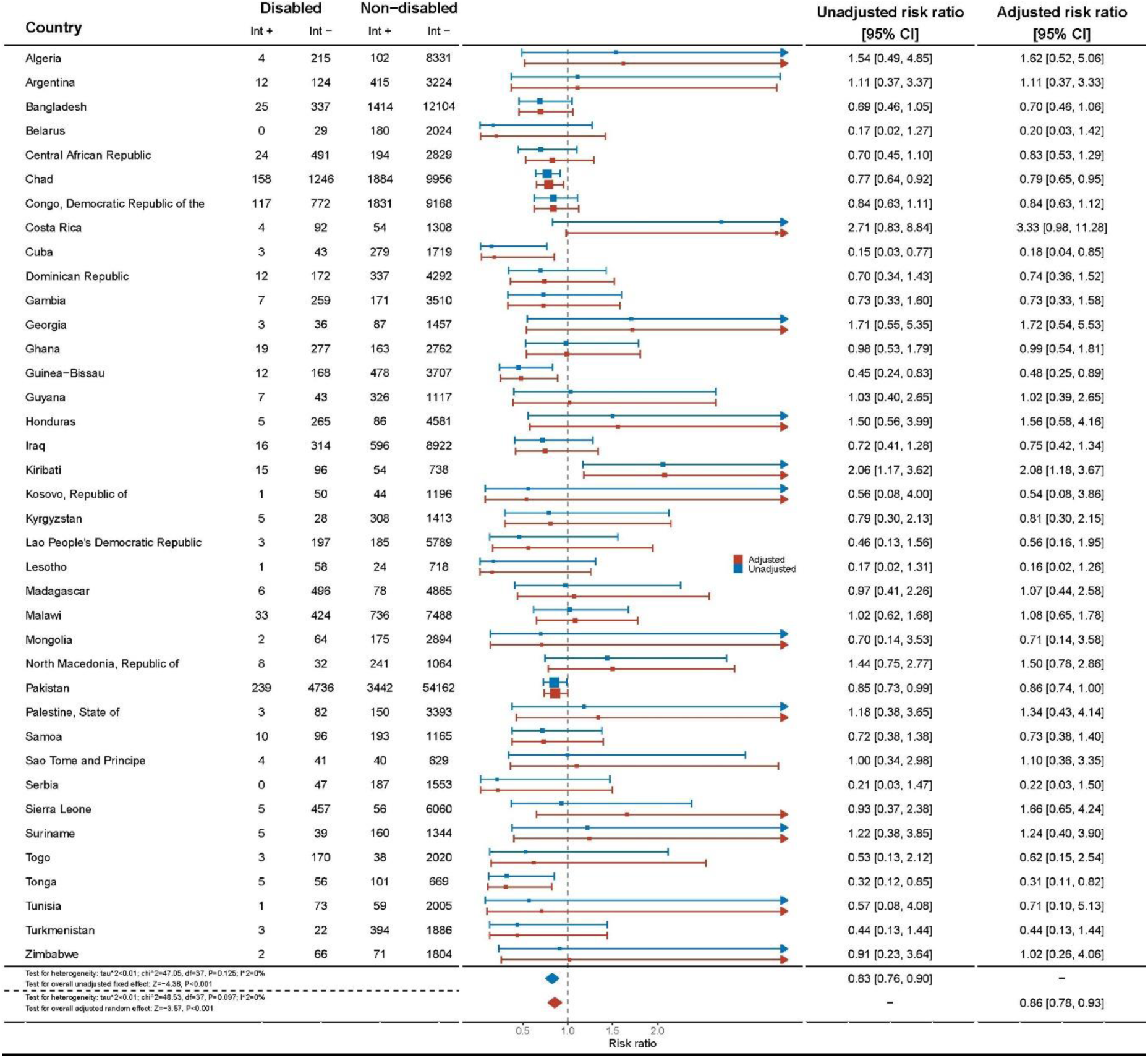

## Appendix 5: Sensitivity analysis

**Sensitivity analysis Table 5.1:**
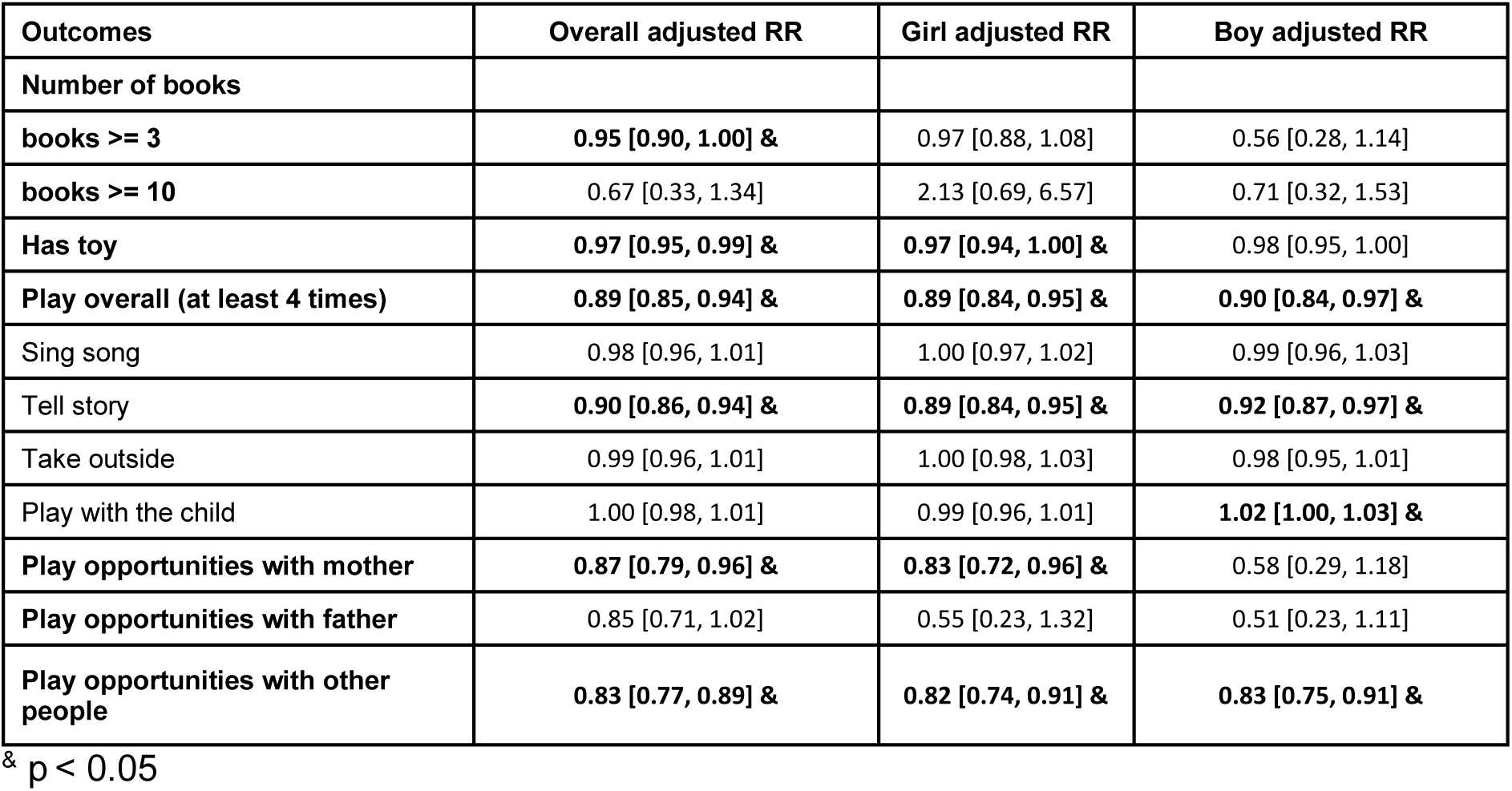
Adjusted risk ratio for play opportunities experienced by children (2-4 years) with disabilities.

**Sensitivity analysis Table 5.2:** Play experiences disaggregated by impairment type (with children without disability as reference)

| impairment | Opportunity to play overall [95%CI] | Mother provides play [95%CI] | Father provides play [95%CI] | Other people provide play [95%CI] |
| --- | --- | --- | --- | --- |
| **Seeing** | 1.14 [0.98, 1.32] | 0.82 [0.45, 1.48] | **2.32 [1.55, 3.51] &** | 0.48 [0.15, 1.52] |
| **Hearing** | 0.62 [0.36, 1.08] | 0.63 [0.28, 1.45] | **3.92 [2.76, 5.62] &** | 1.59 [0.77, 3.29] |
| **Walking** | **0.53 [0.29, 0.99] &** | 0.65 [0.36, 1.17] | 1.17 [0.60, 2.29] | 0.91 [0.47, 1.76] |
| **Fine motor** | **0.41 [0.19, 0.91] &** | 0.56 [0.25, 1.26] | 1.54 [0.61, 3.87] | 0.95 [0.39, 2.31] |
| **Communication** | **0.70 [0.62, 0.78] &** | **0.68 [0.59, 0.80] &** | 0.39 [0.15, 1.00] | **0.37 [0.15, 0.93] &** |
| **Learning** | **0.73 [0.66, 0.81] &** | **0.66 [0.56, 0.79] &** | 0.54 [0.25, 1.15] | **0.49 [0.24, 0.99] &** |
| **Play** | **0.83 [0.73, 0.95] &** | 1.00 [0.85, 1.17] | **1.85 [1.34, 2.55] &** | **1.34 [1.12, 1.81] &** |
| **Behaviour** | 1.04 [0.97, 1.11] | 0.99 [0.88, 1.12] | 1.17 [0.93, 1.47] | 1.07 [0.91, 1.25] |

& p < 0.05

**Sensitivity analysis Figure 5.1:**
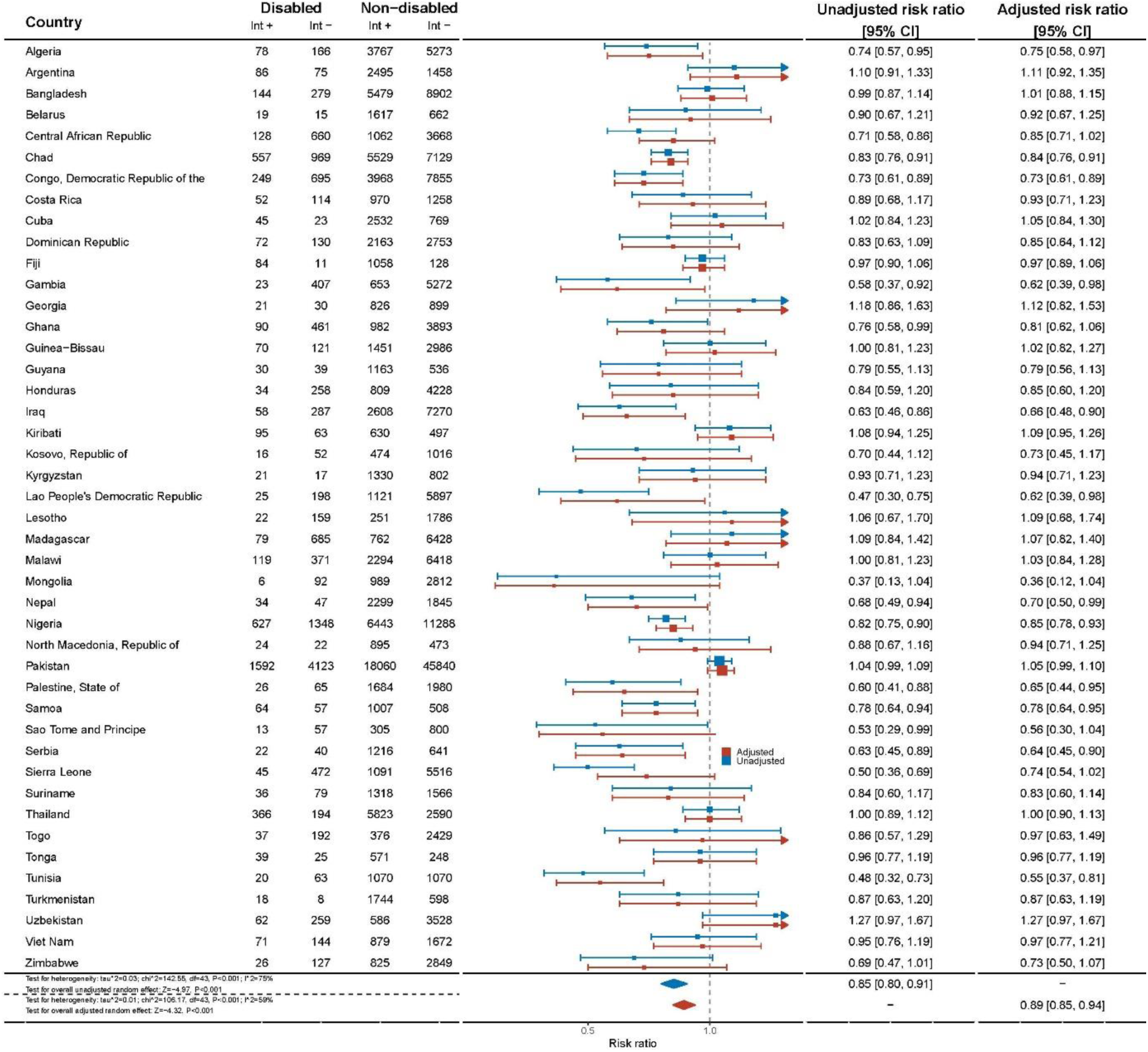
Play opportunities by country*. *Data were extracted from weighted logistic regression models with ‘opportunity to play’ as the outcome and disability status as the key predictor, controlling for age, sex, and wealth status.

**Sensitivity analysis Figure 5.2:**
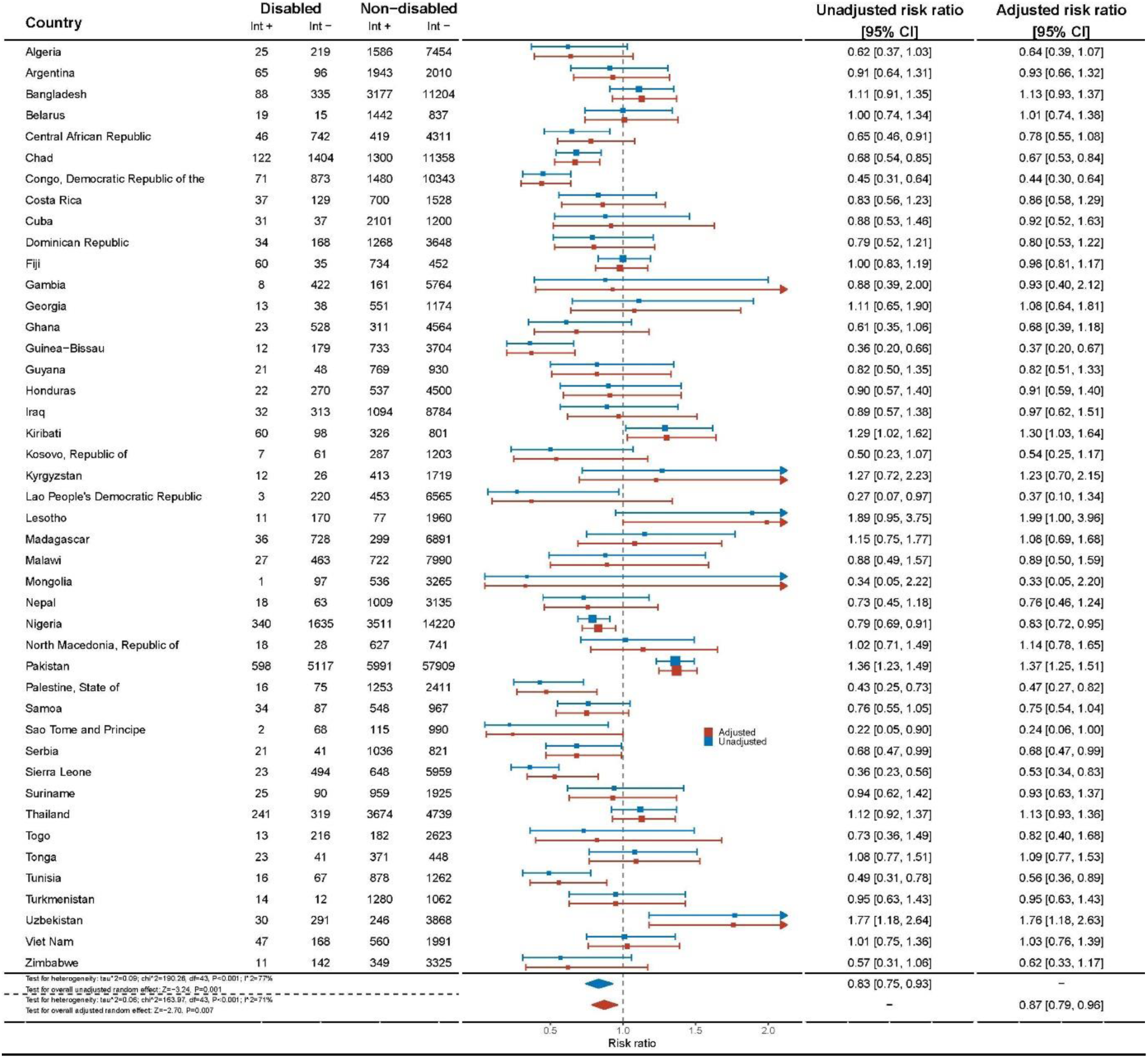
Opportunity to play provided by mother disaggregated by country

**Sensitivity analysis Figure 5.3:**
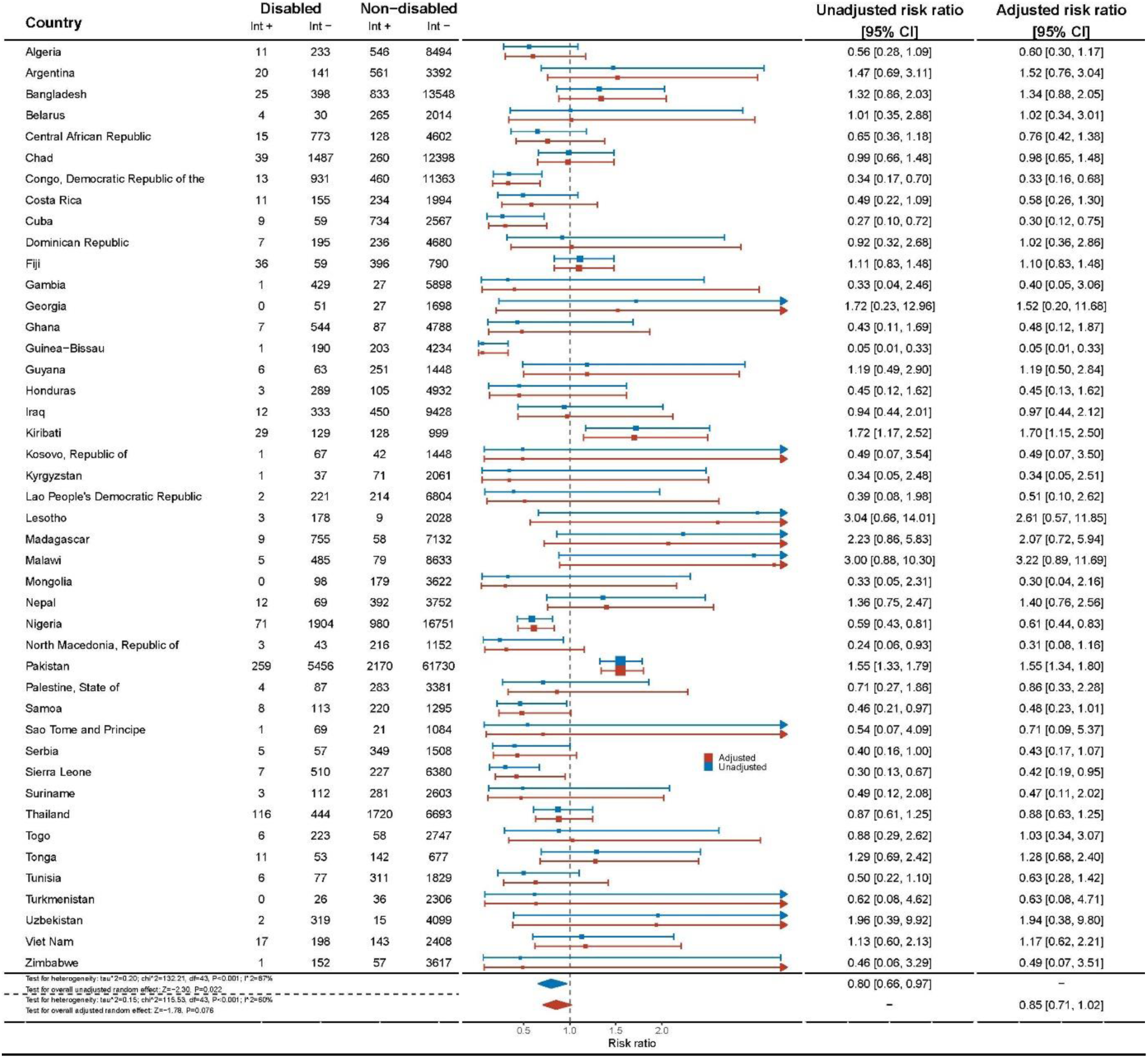
Opportunity to play provided by father disaggregated by country

**Sensitivity analysis Fig 5.4:**
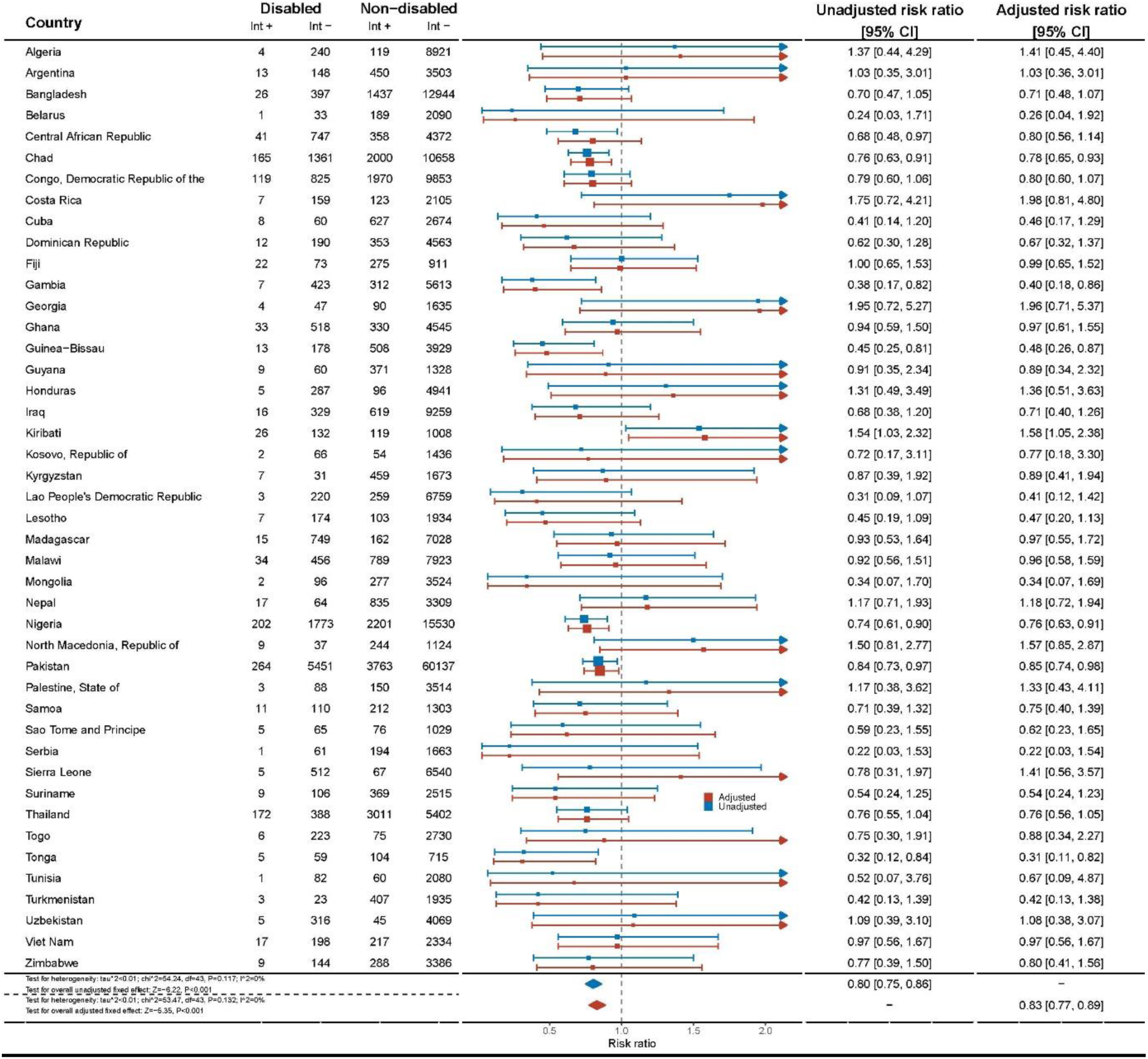
Opportunities for play with other people disaggregated by country

